# Impaired reaching adaptation links to vestibular symptoms

**DOI:** 10.1101/2022.02.07.22270496

**Authors:** Diogo Santos-Pata, Anna Bellmunt, Rosa María San Segundo Mozo, Sandra Sáez, Esther Domènech-Vadillo, Leticia Carballo, Paul F.M.J. Verschure, Belén Rubio Ballester

## Abstract

About 30% of adults suffer from some mild to severe vestibular dysfunction. Vestibular disorders can be expressed as acute vestibular syndrome (AVS), episodic vestibular syndrome. Only half of the sufferers can compensate for their dysfunction after three months after the onset, while the other half of them become chronic, the mechanisms behind this compensation remain unclear. Several behavioural studies have explored the role of the vestibulo-ocular reflex and saccades in the process, linking to the interaction between the superior colliculus and the cerebellum. Yet, despite cerebellar involvement in vestibular function and oculomotor adaptation, thus far, no studies have focused on the specific role of the cerebellum in the compensation process in vestibular dysfunction.

In this study, we test the hypothesis that undiagnosed cerebellar dysfunction might hinder chronic vestibulopathy sufferers from compensating and recovering. We recruited 19 patients who had suffered from an acute peripheral vestibular syndrome (10 clinically recovered and 9 with chronic symptoms) and ten individuals with no history of vestibular alterations (controls). We assessed their implicit motor learning capacity with a visuomotor rotation task and measured by the angular aiming error, which showed an impaired implicit motor adaptation in chronic patients (7.04 ± 1.6º) compared to recovered (11.06 ± 1.94º, *p* = 0.007) and control groups (10.89 ± 7.96º, *p* = 0.03). These findings indicate the involvement of the cerebellum in vestibular compensation and suggest that implicit motor adaptation of reaching movements could be potentially used as an early prognostic tool in unilateral peripheral vestibular dysfunction (UPVD) patients.

## Introduction

About 30% of adults suffer from vestibular dysfunction [1 – 5], increasing to 50% among the elderly [5]. The symptoms, ranging from mild (e.g., dizziness) to severe (e.g., spatial disorientation, disequilibrium, nausea, vomiting), hinder their ability to navigate through the environment, subsequently lowering their independence and quality of life as well as increasing their demands for healthcare services [6].

One of the most prevalent types of vestibulopathy is peripheral and unilateral [2, 46]. Acute unilateral peripheral vestibulopathy (AUPV) is the third most common peripheral vestibular disorder after BPPV and Menière’s disease. The most likely cause is viral (widely called vestibular neuritis), although it also may result from other infections, trauma, or postoperative complication. Given that clinicians cannot visualize vestibular structures, the primary diagnosis relies on behavioral manifestations of the pathology, such as an impaired vestibulo-ocular reflex (VOR) [8]. After ruling out central structural damage in the brainstem and cerebellum using imaging techniques [9], patients are diagnosed with peripheral origin. The prognosis of peripheral vestibulopathies is varied: after three months post-onset, half of the patients continue to experience impairing symptoms even after treatment (chronic patients) [9-15], while others can compensate or adapt for their dysfunction and return to their everyday lives (remission).

The reason behind the patients’ ability to recover - or lack thereof - remains unclear. Several studies have tried to identify it by exploring the correlation between symptomatology and oculomotor mechanisms (i.e., VOR and saccades) using the video Head Impulse Test (vHIT) [4, 16-17]. However, their findings are not conclusive; although abnormal VOR gain values indicate dysfunction [18, 19], some patients show abnormal VOR patterns while being in remission and vice versa [17, 20-21].

One aspect which questions the effectiveness of oculomotor mechanisms as a diagnostic tool is that they are not exclusively related to the vestibular system. Other structures could modulate the contribution of these mechanisms to vestibular compensation and VOR adaptation. For instance, the cerebellum is structurally and functionally linked to the vestibular system [10, 26, 45], and it is of central importance in controlling eye movements [26, 27] and also in VOR gain and implicit motor adaptation [28-33]. However, its specific role in UPVD is not fully understood. UPVD symptoms are often attributed to peripheral processes (Hess et al. 2000); however, an animal study with impaired mice unable to increase oculomotor gain after vestibular damage demonstrated that vestibular compensation requires functional olivo-cerebellar circuitry, proposing that UPVD recovery may largely depend on central rather than peripheral adaptive mechanisms [33b]. Following this line of research, here we assess whether undiagnosed functional cerebellar alterations can explain the absence of spontaneous recovery in humans with UPVD.

Implicit motor adaptation of reaching movements has proven helpful as a behavioural indicator of cerebellar function and is frequently studied with visuomotor rotation (VMR) paradigms [34, 35]. In VMR tasks, subjects experience a mismatch between a performed motor action and the observed outcome, which creates a conflict between the predicted trajectory and the actual trajectory. Individuals without neurological damage gradually modify their executed motor action to compensate for the mismatch, a behavior often called implicit motor adaptation [36, 37]. However, participants with structural cerebellar damage [38, 39] or functional cerebellar alterations [39b, Herreros, et al., 2019] display reduced implicit adaptation.

To explore possible alterations in cerebellar function in vestibulopathy patients, we tested a VMR task derived from Taylor et al. [34], in both chronic and recovered patients (Figure 1). This task has been repetitively used in previous studies to probe a putatively-cerebellar implicit motor adaptation process together with the learning and execution of an explicit aiming rule (Herreros, et al., 2019; Schween, et al., 2014). We hypothesize that an undiagnosed olivo-cerebellar dysfunction might be hindering chronic vestibular patients from spontaneously developing compensatory oculomotor mechanisms, and lower directional implicit adaptation rates can diagnose this malfunction during arm reaching.

**Figure 1.**
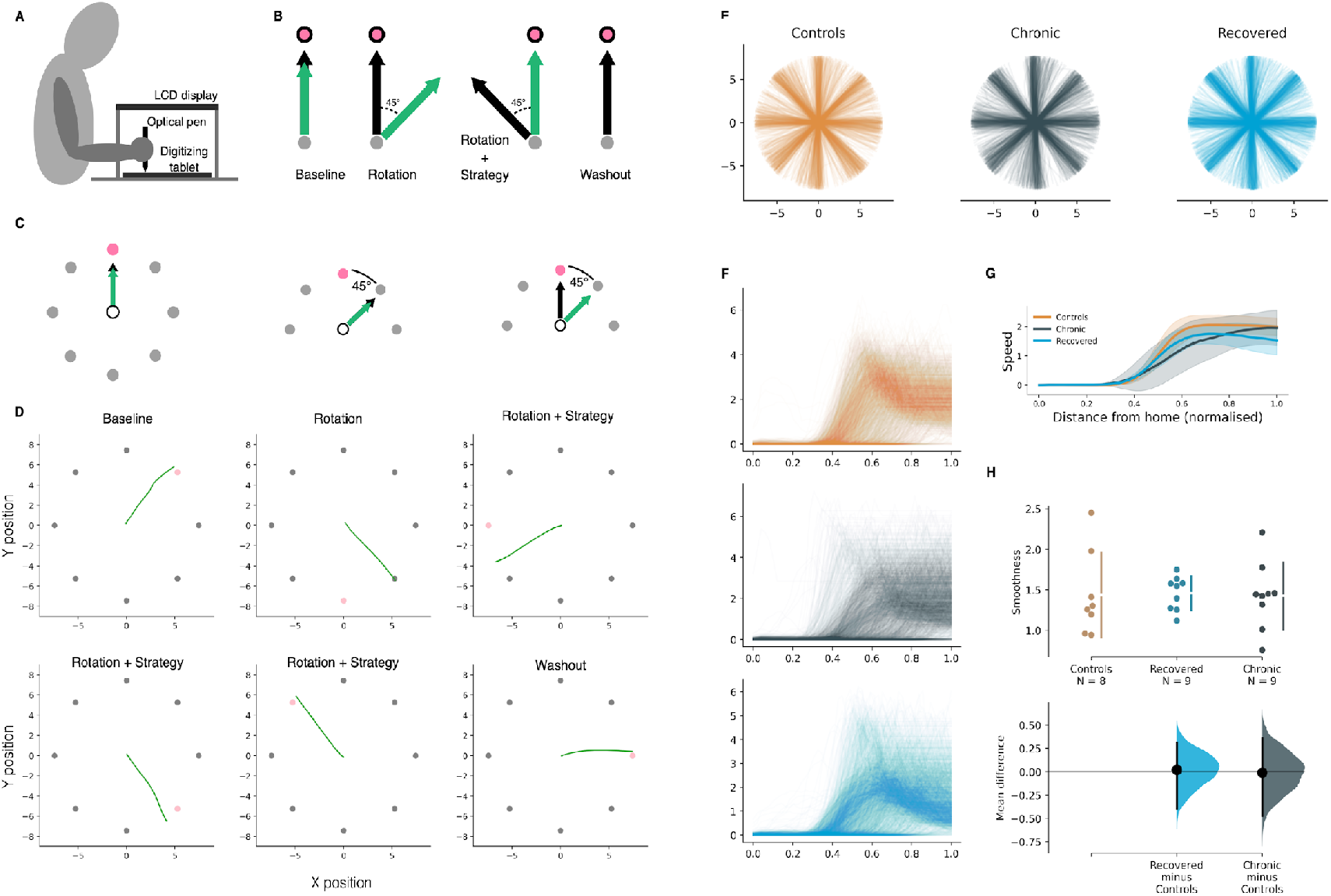
VMR task setup and phases. (**A**): Configuration of the experimental setup. Subjects were seated in front of a table and looked at a 15-inch screen mounted 25.4 cm above it. Data were sampled and stimuli displayed at 100 Hz; movement trajectories were recorded through a digitizing tablet (Intuos Pro, Wacom, Saitama) located at waist height at the lower part of the table below the screen. (**B**): Visuomotor task: Baseline, Rotation, Rotation+Strategy and Washout. The cursor (green arrow) reproduces hand movement (black arrow). **(C):** (Left) A centered home position (white circle) and eight circles are displayed around the home position. At the start of each trial, a bullseye appears in one of the eight circles, indicating the aiming target. (Middle) In the Strategy training phase, the subjects are instructed to aim at the circle adjacent to the target (i.e., the neighbour target, pink circle). (Right) In the Rotation phase, we introduce the rotation bias, the cursor’s movement is rotated by 45 degrees in a counter-clockwise direction, and subjects are instructed to aim at the circle adjacent to the target (i.e., the neighbour target, pink circle) to compensate for the directional bias. **(D):** Sample cursor trajectories per task phase. The pink circle indicates the neighbour target. **(F):** Speed profiles for all trials executed by all subjects per group. **(G):** Averaged speed profiles per group. Gray shaded areas indicate standard deviations. **(H):** Comparison of the curvature of hand trajectories across groups.

**Figure 2.**
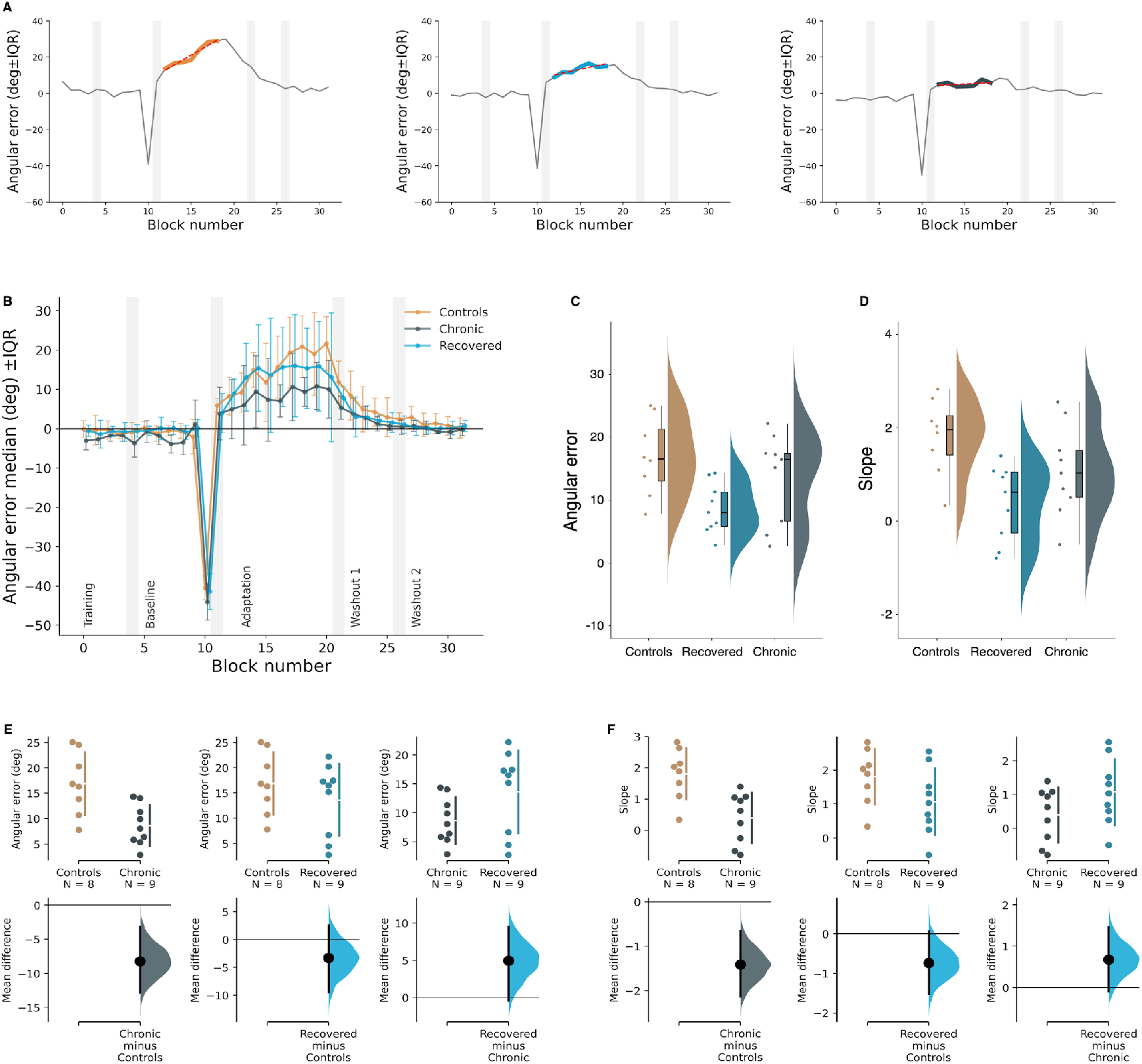
Implicit motor adaptation. (**A**): Slope fit during the Rotation+Strategy phase for one representative subject per group. **(B):** Directional aiming errors throughout the phases of the VMR task. Vertical shaded areas indicate the first 8-trials bin of each phase. The horizontal line shows the target direction (i.e., no directional error, 0° error). Each point represents the median of the errors of each block of 8 trials. Error bars indicate IQRs. (**C-D**): Group comparison of median angular errors and regression line fits during the Rotation+Strategy phase. (**E-F**): Pairwise comparisons of median of errors and regression line fits during Rotation+Strategy phase. *: p < 0.05; **: p < 0.01; NS: not significant. Error bars indicate the IQR.

## Results

### Sample demographics

Chronic patients (n = 9, 3 women) had a mean age of 55.3 ± 12.9 years old, which was age-matched by the recovered group of patients (n = 9, 3 women; 57.7 ± 6.6 years old) (Table 1). Chronic patients’ dysfunction lasted more than three months, with remaining impairing symptoms; recovered patients did not show any symptoms. In addition, we recruited a control group (n = 9, 6 women, 34.5 ± 14.3 years old) of individuals with no diagnosed conditions.

**Table 1.** Demographic and clinical characteristics of the experimental groups. VOR: vestibulo-ocular reflex.

| | Chronic ($n = 9$) | | | | Recovered ($n = 9$) | | | | *p*-value |
| --- | --- | --- | --- | --- | --- | --- | --- | --- | --- |
|  | *n* | % | Mean | SD | *n* | % | Mean | SD |  |
| **Age** |  |  | 55.3 | 12.9 |  |  | 50.7 | 11.1 | 0.51 |
| **Sex** |  |  |  |  |  |  |  |  |  |
| Male | 4 | 66.6 |  |  | 6 | 66.6 |  |  |  |
| Female | 2 | 33.3 |  |  | 3 | 33.3 |  |  |  |
| **Etiology** |  |  |  |  |  |  |  |  |  |
| Vestibular Neuritis | 2 | 33.3 |  |  | 9 | 100 |  |  |  |
| Post-operative | 4 | 66.6 |  |  | 0 | 0 |  |  |  |
| **Affected side** |  |  |  |  |  |  |  |  |  |
| Right | 5 | 83.3 |  |  | 4 | 44.4 |  |  |  |
| Left | 1 | 16.6 |  |  | 5 | 55.5 |  |  |  |
| **VOR gain** |  |  | 0.65 | 0.08 |  |  |  |  |  |
| **Saccades** |  |  |  |  |  |  |  |  |  |
| Covert and overt | 2 | 33.3 |  |  |  |  |  |  |  |
| Overt | 4 | 66.6 |  |  |  |  |  |  |  |

SD: Standard Deviation

### Between-groups differences in implicit motor adaptation

We first analyzed the evolution of the angular error during the VMR task. Notice that we arranged the task (Figure 1 A-C) in different phases that showed eight circles around a central home. At the start of each trial, a bullseye appeared randomly in one of the peripheral circles (target). We instructed the subjects to perform a straight line from the home position towards the target to overshoot its location and return to the center. In the experimental phase (Rotation + Strategy; RS), cursor trajectories were rotated, and subjects were told to counteract the rotation. We extracted aiming errors during the RS phase [40] together with fits of the slope of the regression line for each group to quantify implicit adaptation and compare it across groups. All measures were baseline normalized.

After the onset of the RS phase, we found adaptation occurred early in all the groups, as has been shown in other studies with continuous feedback [41] (Figure 1 D-E). There were no differences between groups in the speed profiles (Kurskal-Wallis, *p*>0.1, Figure 1 F-G) and the curvature (Kurskal-Wallis, *p*>0.1, Figure 1 H) of hand trajectories.

The analysis of errors during the adaptation phase revealed a main effect of the group (Kurskal-Wallis, H(2)= 6.44, *p*=0.04). Pairwise comparisons showed the main group effect was due to a significant difference between chronic patients (mean 8.66 ± 4.00º std) and individuals without diagnosed conditions (mean 16.90 ± 6.18º std, Mann Whitney U=62, p = 0.01). We observed no differences in implicit motor adaptation between recovered (mean 13.61 ± 7.13º std) and control groups (mean 16.90 ± 6.18º std, Mann Whitney U=44, *p* = 0.60). Results from the fitted slopes displayed consistent results, showing a main effect (Kurskal-Wallis, H(2)= 8.66, *p*=0.01) and a significant difference in implicit adaptation between chronic (mean 0.40 ± 0.81º std) and control groups (mean 1.81 ± 0.81º std, Mann Whitney U=66, *p*<0.01).

All groups performed similarly when compared amongst themselves at baseline (mean -1.54 ± 1.40º std for chronic, mean -0.38 ± 1.25º std for recovered, and mean -0.61 ± 0.89º std for controls, Mann Whitney U, *p* > 0.05).

## Discussion

In this study, we investigated the question of whether recovering and non-recovering UPVD patients might be distinguished in their implicit learning capabilities. This hypothesis is based on the consideration that vestibular deficits and their symptomatology, such as deficits in VOR, pertain to a broad network of neuronal systems, including the superior colliculus and cerebellum, where the latter plays a key role in the compensation of vestibular behaviors. Using an adaptation of a visuomotor rotation task that looks at the interaction between explicit rule-based and implicit procedural learning systems, we show that non-recovering patients display significant impairments in implicit motor adaptation. Hence, this suggests that a finer-grained triage of UPVD patients is required, taking cerebellum-dependent implicit learning into account. In addition, our task provides a straightforward diagnostic test to distinguish between recoverers and non-recoverers.

Chronic vestibular patients frequently refer to healthcare systems [6] and present comorbidity with disorders such as depression or anxiety [43, 44]. Theirs is an incapacitating, isolating, and uncertain disorder. There is still no way to predict which patient who suffers from an acute peripheral vestibulopathy is at risk of becoming a chronic patient. Our study presents a plausible explanation that can account for the differences in oculomotor manifestations and in the symptomatology of recoverers and non-recoverers (i.e., chronic patients). The visuomotor adaptation paradigm we applied has been successfully used as a marker of cerebellar damage in ataxic patients [34]. Like ataxic patients, chronic vestibulopathy patients displayed significantly fewer errors than recovered patients and controls, indicating an impaired implicit motor adaptation. These results are consistent with our hypothesis that a malfunction of the olivo-cerebellar circuit, although not clinically symptomatic, might be preventing chronic patients from compensating for their vestibular dysfunction. Further research should assess patients’ implicit motor learning when they first refer to the clinic with acute symptoms and follow their evolution with a longitudinal study to discriminate whether cerebellar malfunction precedes or follows their vestibulopathy.

## Conclusions

Contrarily to patients with unilateral peripheral vestibulopathies who recover within 3 months from the dysfunction’s onset (recoverers), those with chronic symptoms show an impaired capacity for implicit motor learning during arm reaching. Therefore, chronic patients may suffer from an undiagnosed olivo-cerebellar malfunction that compromises the mechanisms involved in compensating their vestibular impairment.

## Methods

### Demographic information

An otorhinolaryngology specialist from Hospital Joan XXIII recruited patients who referred to their consultation due to vestibular symptoms at the time of the study, or had consulted them in the past, and met the following inclusion criteria:

– > 18 years old.
– Diagnosed with one of the following UPVD: vestibular neuritis, sudden hypoacusis with vestibular affectation, labyrinthitis, Ménière’s disease, vestibular schwannoma.
– Presented vestibular symptoms (vertigo, dizziness, disequilibrium, walking instability, kinetosis, or oscillopsia) or had suffered from them in the past three years.
– The onset of the dysfunction, or the symptoms, had occurred ≥ 3 months prior.

Patients who met any of the following exclusion criteria were excluded:

– Diagnosed neurological, traumatological, rheumatological, ophthalmological, or systemic pathology that could interfere with equilibrium.
– Diagnosed benign paroxysmal positional vertigo.
– Not understanding the participation in the study or refusing to participate.
– Not meeting any of the inclusion criteria.

17 patients met the inclusion criteria (mean age = 52.6, SD = 12.1): 12 patients were diagnosed with vestibular neuritis, 1 with Ménière’s syndrome, and 4 with symptoms due to post-operation complications.

According to their evolution, patients were divided into two groups: recovered and chronic. Chronic patients reported symptoms at the time of the study and manifested them when evaluated with a series of clinical tests led by an otorhinolaryngology specialist, who also collected socio-demographic data, including gender and age. Recovered patients did not report any symptoms.

We also recruited 11 individuals without vestibular alterations (mean age = 33.6, SD = 13.9).

The protocol was approved by the review board of the Institut d’Investigació Sanitària Pere Virgili (Comitè Étic d’Investigació amb medicaments), and all participants provided informed consent before the experiment.

### Clinical tests

The patients completed a series of clinical tests at recruitment and the day they participated in the study, led by a rehabilitation or otorhinolaryngology specialist. The tests performed (vHIT, Romberg Lab, Standing Balance, Timed Up and Go, Tinetti, Barthel, and SF36) assessed their affectation and symptomatology.

### Equipment

Participants sat in front of a desk, with the vision of their hand occluded by a monitor. Visual feedback was provided in a small cursor (3.5 mm), displayed on the screen, and controlled by an optical pen. Movement trajectories were sampled at 100 Hz using a high-resolution digitizing tablet (Intuos3, Wacom, Saitama, Japan). The stimuli and feedback cursor were displayed on a 15 inch, 1280 × 1024-pixel resolution LCD portable computer and horizontally mounted 25.4 cm above the tablet.

### Experimental Procedure and Conditions

During the task, subjects were presented on the monitor with eight circles separated by 45°. Every trial required subjects to perform unimanual reaching movements with their right. At the start of each trial, a bullseye appeared randomly inside one of the eight circles (target). Participants were instructed to perform a center-out, overshooting movement towards it by sliding an optical pen across a tablet. They were asked to be fast, move straight, and overshoot the target location, avoiding online corrections during the movement execution. To force fast-reaching movements, trials were repeated when movements were not completed within a time limit of 1.5 seconds after the target appearance. When the subject performed the movement correctly, the target turned green, and they had to return to the home position.

In total, subjects underwent a total of 242 movements or trials, divided into 6 phases. These movements were grouped in 31 blocks of 8 trials and 1 block of only 2 trials. We replicated the protocol performed in cerebellar ataxic patients [34]. The first and second phases were introduced for training and familiarization purposes. In the first phase (Practice Baseline) the participants were instructed to aim at the target location. In the second phase (Practice Strategy), they were asked to apply an explicit strategy and aim at the circle adjacent to the target, deliberately producing a 45° clockwise error. During both phases, the cursor reproduced the participants’ hand movement, and participants were encouraged to ask any doubts about the execution of the task. In the third phase (Baseline) participants were again instructed to aim at the target location. In the fourth phase (Rotation), subjects did not get any instructions, and for two consecutive trials the hand movement to cursor mapping was rotated -45° relative to their hand movement (counter-clockwise). After these two trials, subjects were instructed to counteract the imposed rotation in the fifth phase (Rotation + Strategy) by applying the same strategy they used in the Practice Strategy phase. Finally, in the sixth phase (Washout), visual feedback was reinstalled in the absence of any rotation to test for aftereffects. The whole task lasted approximately 20 minutes.

### Quantification and statistical analysis

We measured performance in angular error (i. e., angular difference between the executed movement and target position). To account for individual trial noise, we calculated the angular error per block of 8 trials each. In each of these blocks, all eight target positions were presented once. Average and dispersion values are reported in medians ± IQR.

For all tests, statistical significance levels were set at P < 0.05. All statistical analyses were performed using Python 3.6.

## Data Availability

All data produced in the present study are available upon reasonable request to the authors

## Data availability

At the time of publication, all data used in this study will be freely available at OSF. Any additional information concerning the data used in this study will be made available from the corresponding author upon reasonable request, provided this information can be made available in anonymized form. Any additional data and information are available from the corresponding author on reasonable request.

## Code availability

At the time of publication, this study’s code and related data will be freely available at OSF, enabling reproduction.

## Acknowledgements

The authors gratefully acknowledge the participation of the subjects. This study was supported by the European Commission (EC), the European Research Council under grant agreement 341196 (CDAC), EC H2020 project socSMCs (H2020EU.1.2.2. 641321), and by the RGS@home, EIT Health project 19277. EIT Health is supported by EIT, a body of the European Union.

## Author contributions

B.R.B and D.S.P. conceptualized the analysis; B.R.B, R.m.S.S.M, S.S., E.D.V., L.C, and P.F.M.J.V. designed the methodology; R.m.S.S.M, S.S., E.D.V., and L.C constructed the datasets; B.R.B, and D.S.P.. performed data analysis; and B.R.B, D.S.P., R.m.S.S.M, S.S., E.D.V., L.C, and P.F.M.J.V. wrote the manuscript.

